# Genetic analysis implicates *ERAP1* and HLA as risk factors for severe Puumala virus infection

**DOI:** 10.1101/2024.06.28.24309633

**Authors:** Hele Haapaniemi, Satu Strausz, Anniina Tervi, FinnGen, Samuel E. Jones, Mari Kanerva, Anne-Marie Fors-Connolly, Hanna M. Ollila

## Abstract

Puumala virus (PUUV) infections can cause severe illnesses such as Hemorrhagic Fever with Renal Syndrome in humans. However, human genetic risk factors contributing to disease severity are still poorly understood. Our goal was to elucidate genetic factors contributing to PUUV infections and understand the biological mechanisms underlying individual vulnerability to the disease. Leveraging data from the FinnGen study, we conducted a genome-wide association study on severe Hemorrhagic Fever with Renal Syndrome caused by PUUV with 2,227 cases. We identified associations at the Human Leukocyte Antigen (HLA) locus and *ERAP1* with severe PUUV infection. HLA molecules are canonical mediators for immune recognition and response. *ERAP1* facilitates immune system recognition and activation by cleaving viral proteins into smaller peptides which are presented to the immune system via HLA class I molecules. Notably, we identified that the lead variant (rs26653, OR = 0.84, p = 2.93×10-8) in the *ERAP1* gene was a missense variant changing amino acid arginine to proline. From the HLA region, we showed independent and significant associations with both HLA class I and II genes. Furthermore, we showed independent associations with nine HLA alleles and severe PUUV infection using conditional HLA fine-mapping. The strongest association was found with the *HLA-C*07:01* allele (OR = 1.5, p = 4.0×10^−24^) followed by signals at *HLA-B*13:02, HLA-DRB1*01:01*, and *HLA-DRB1*11:01* alleles (p<5×10^−8^). Our findings suggest that viral peptide processing with *ERAP1* and antigen presentation through HLA alleles contribute to the development of severe PUUV disease.

## Introduction

Puumala orthohantaviruses (PUUV) belong to the Hantaviridae family. PUUV primarily circulates among rodent populations establishing persistent infections and transmits to humans through contact with rodent excretions. (1) Of the two hantaviral diseases, Hantavirus Pulmonary Syndrome (HPS) and Hemorrhagic Fever with Renal Syndrome (HFRS), PUUV causes only HFRS which is characterized by symptoms such as fever, kidney dysfunction, and hemorrhagic complications. Head and back pain are common symptoms, and chest X-ray findings and transient ocular myopia may also occur during the acute stage of PUUV. Despite having a low fatality rate, approximately 52% of the symptomatic cases require hospitalization (2) and 5 % of the hospitalized patients need dialysis or intensive care treatment for acute kidney injury (AKI) (1). In addition, PUUV often results in complications and long-term hormonal, renal or cardiovascular consequences. (3)

In Finland, PUUV has a significant impact on public health. The reported average incidence rate of diagnosed PUUV infections in Finland is 31-39 cases per 100,000 inhabitants. (4) However, the true incidence rate is considered to be seven to eight times higher based on the seroprevalence rate (12.5% in Finland). (5)

Due to the high incidence of PUUV infections in Finland, much research has been invested in epidemiology, virology, clinical courses, and the ecology of the virus in the carrier bank vole (Myodes Glareolus). The effect of host genetics on PUUV disease susceptibility and severity has primarily been explored through Human Leukocyte Antigen (HLA) haplotype studies. Small case-control studies have identified several HLA alleles and haplotypes associated with severe disease, although the findings vary between studies. (6-9)

We utilized data from 520,210 individuals participating in the FinnGen project to explore the biological mechanisms behind PUUV infection. With International Classification of Diseases (ICD) code-based case definition, we identified 3,650 cases of PUUV infection diagnosed between 1995 and 2023. Out of these cases, 2,227 required hospitalization and were classified as severe. We assessed the host genetic component with Genome-Wide Association study (GWAS) and fine-mapping of the HLA region and explored the comorbidity burden with epidemiological analyses.

## Results

### PUUV infection is associated with thrombocytopenia and acute renal failure

Using data from FinnGen, we identified 3,650 individuals (8.3%) with ICD10 code A98.5-based Hemorrhagic fever with renal syndrome. In Finland, these cases are primarily caused by Puumala hantavirus. We first performed a descriptive analysis of the cohort to understand the associations between PUUV infection and demographic factors. We observed the highest incidence of PUUV infections among people aged 50-60 years (median 52 years) (Figure 1B). In addition, we saw variation in incidence rates within a year as well as between years. The highest incidence rates were seen in autumn and a peak year occurs every 3-4 years following the changes in vole population as reported previously (10) (Figure 1A and 1C). Lastly, we observed an over-representation of males within the cases (54.0 %) compared to controls (43.5 %) (p = 2.4×10^−6^) and a slightly higher BMI in cases (27.7) versus controls (27.3) (t = 3.3, p = 8.8×10^−4^).

**Figure 1.**
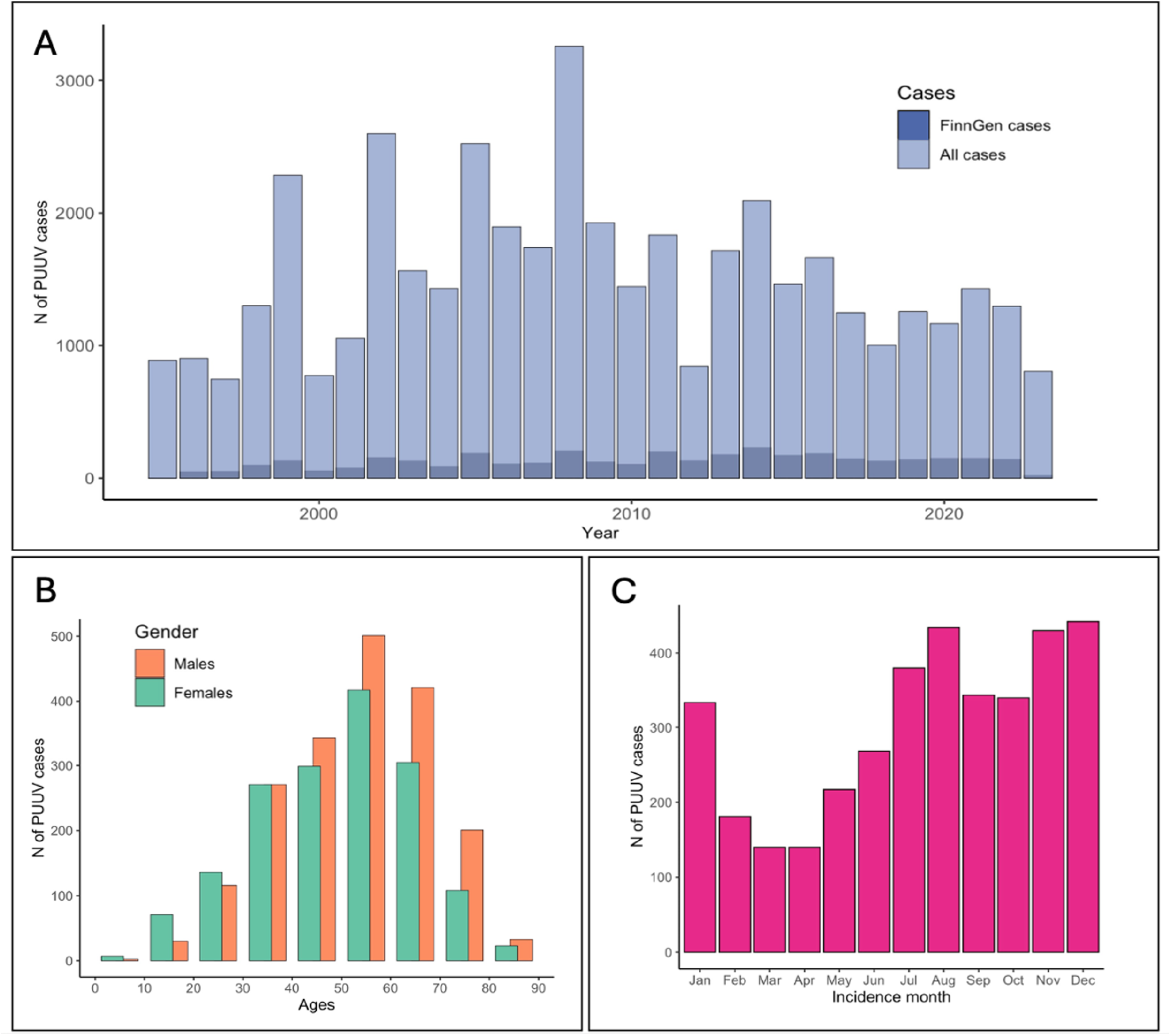
A) Annual numbers of Puumala virus (PUUV) cases by ICD-10 coding in FinnGen and total numbers of new PUUV diagnoses notified by laboratories in the National infectious diseases registry in Finland. FinnGen has data on about half a million Finns, which corresponds to almost 10 % of the total population in Finland. Incidences for all Finns were obtained from the Finnish National Infectious Diseases Register. B) Age at the first PUUV diagnosis in FinnGen for males and females. C) Monthly incidence of PUUV in FinnGen.

Previous research studies and case reports have documented an elevated occurrence of thrombocytopenia (11), acute renal failure (11), thrombosis (11), lymphoid malignancies (12), and hypopituitarism (13) after PUUV infection. We employed multivariate logistic regression to study the possible association of PUUV and the conditions previously associated with it. We report association with with thrombocytopenia (OR = 2.46 [1.84, 3.30], p = 5.14×10^−8^), acute renal failure (OR = 1.31 [1.27, 1.35], p = 4.33×10^−6^), hypopituitarism (OR = 1.69 [1.39,2.05], p = 6.50×10^−3^), lymphoid malignancies (OR = 1.33 [1.24, 1.42], p = 0.019) and thrombosis (OR = 1.24 [1.19, 1.28], p = 0.018) overall as complications of Hantavirus infection (Table S2). Subsequently we employed Cox proportional-hazards models to study the temporal association of the conditions (Table S3). Associations of PUUV with thrombocytopenia (HR = 1.86 [1.34, 2.58], p = 2.22×10^−4^) and acute renal failure (HR = 1.22 [1.10, 1.34], p = 9.87×10^−5^) outcomes were supported by the Cox proportional-hazards models. In addition, temporal association with thrombosis was significant although negative (HR = 0.76 [0.63, 0.92], p = 4.58×10^−3^).

### A genome-wide association study highlights *ERAP1* and *HLA* class I and II loci

To investigate the role of single nucleotide polymorphisms (SNPs) in severe PUUV infection, we conducted a genome-wide association study (GWAS) using a dataset comprising 3,650 individuals diagnosed with Haemorrhagic fever with renal syndrome (ICD A98.5) sourced from FinnGen Release 12. Of these cases, 2,227 (61%) were admitted to hospital and grouped to severe cases whereas 1,423 were classified as non-severe cases. We performed two GWAS analyses for severe cases, one using the remaining participants in the FinnGen cohort (= population control, N = 498,175) and another using non-severe HFRS cases as controls.

We identified genetic variants at the *ERAP1* gene in chromosome 5 and the HLA region in chromosome 6 with genome-wide significant associations (p < 5×10^−8^) with severe PUUV infection using population control (Figure 2, Figure S1, Figure S3, Table S4). The lead variant in chromosome 5 was a missense variant in the second exon of the *ERAP1* gene (rs26653, p = 2.93×10^-8,^ beta = -0.18, AF = 0.71) changing amino acid arginine in position 127 to proline.

**Figure 2.**
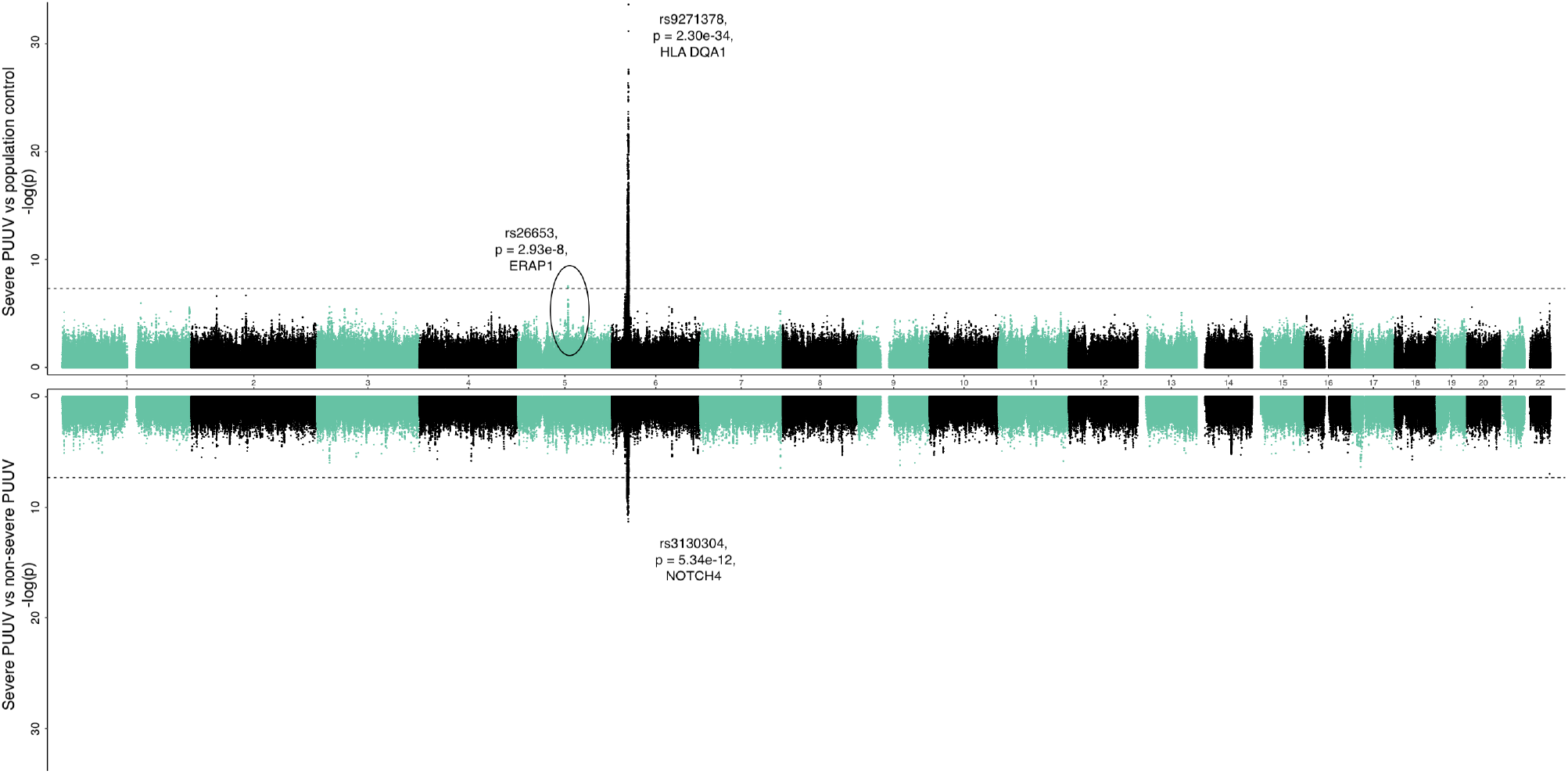
Miami plot for severe hemorrhagic fever with renal syndrome due to Puumala virus (PUUV) using population control (upper, N = 498,175) and mild cases as controls (lower, N = 1,423). The X-axis represents the chromosomal position for each variant. The Y-axis shows the -log10(P)-value. The horizontal line indicates the genome-wide significance threshold of p = 5×10^−8^.

The lead variant in chromosome 6 (rs9271378, p = 2.30×10^−34^, beta = 0.36, AF = 0.41) was located at the HLA class II region between *HLA-DRB1* and *HLA-DQA1*. Additionally, conditional GWAS analysis adjusting with the HLA class II lead variant, rs9271378, showed a second genome-wide significant independent association at the HLA class I region closest to the *HLA-C* gene (rs2844614, p = 1.0×10^−13^, beta = 0.31, AF = 0.13).

To explore if the same genetic factors contribute to non-severe PUUV infection and severe PUUV infection requiring hospital treatment, we compared severe PUUV cases to non-severe PUUV cases. The non-severe cases had a laboratory-confirmed PUUV infection but did not require hospital treatment. Similar to the severe PUUV versus population control GWAS analysis, we identified a genome-wide signal at the HLA region (Figure 2, Figure S2-S3, Table S3). The lead variant was closest to *NOTCH4* gene (rs3130304, p = 5.34×10^−12^, beta = 0.46, AF = 0.19), and located approximately 200 kilobases from the HLA class II region. We also observed an association with *ERAP1* lead variant rs26653 (p = 8.3×10^−5^, beta = -0.22).

The genome-wide significant association with both *ERAP1* and HLA region highlights the role of viral peptide processing and presentation in defense against PUUV.

### HLA fine-mapping identifies the strongest association with *HLA-C*07:01*

The HLA region is known for its high genetic diversity and many variants are in strong linkage disequilibrium (LD) with each other. To understand the complexity of genetic associations, identify the specific causal HLA alleles, and gain a deeper understanding of the biological mechanisms influencing the PUUV infection, we performed fine-mapping of the HLA region.

We used imputed HLA allele information to assess whether HLA alleles were associated with susceptibility to PUUV. We included alleles with minor allele frequency greater than 1% in the fine-mapping analysis (Supplementary Excel Table 1). We used multivariate logistic regression and adjusted the analysis for age at death or the end of follow-up, sex, and the first 10 genetic principal components. We found 46 significant associations with HLA alleles (p < 0.05). Due to high linkage disequilibrium (LD) in the HLA region, not all associations were expected to be independent. We performed a stepwise logistic regression model to identify the individually associated HLA alleles, adjusting for the most strongly associated HLA allele until the lead variant was not statistically significant (p ≥ 0.05). We found nine independent HLA alleles associated with severe PUUV (using population control) out of which eight alleles were predisposing and one protective (Table 1, Figure 3A-B, Supplementary Excel Table 2). Using hospitalized individuals as cases and non-hospitalized individuals as controls, we identified 25 significant associations at the first fine-mapping round out of which nine were independent with five predisposing and three protective alleles (Table 2, Figure 3C-D, Supplementary Excel Table 3).

**Table 1.** Associations of independent HLA alleles to severe Puumala virus (PUUV) infection were analyzed using fine-mapping and stepwise logistic regression. Individuals with severe PUUV were used as cases and individuals without hospital level diagnosis of PUUV infection were classified as controls. (se = standard error, carrier frequency = frequency of having at least one copy of allele).

| HLA allele | p | beta | se | carrier frequency | Alleles used for adjusting |
| --- | --- | --- | --- | --- | --- |
| *C\*07:01* | 4.0E-24 | 0.43 | 0.04 | 0.23 | NA |
| *B\*13:02* | 1.2E-17 | 0.61 | 0.07 | 0.06 | *C\*07:01* |
| *DRB1\*01:01* | 1.5E-14 | 0.30 | 0.04 | 0.33 | *C\*07:01, B\*13:02* |
| *DRB1\*11:01* | 4.1E-17 | 0.66 | 0.08 | 0.07 | *C\*07:01, B\*13:02, DRB1\*01:01* |
| *C\*07:04* | 4.3E-05 | 0.40 | 0.10 | 0.04 | *C\*07:01, B\*13:02, DRB1\*01:01, DRB1\*11:01* |
| *DQB1\*02:02* | 2.8E-04 | 0.36 | 0.10 | 0.08 | *C\*07:01, B\*13:02, DRB1\*01:01, DRB1\*11:01, C\*07:04* |
| *DRB5\*01:01* | 9.6E-04 | -0.17 | 0.05 | 0.26 | *C\*07:01, B\*13:02, DRB1\*01:01, DRB1\*11:01, C\*07:04, DQB1\*02:02* |
| *DRB1\*09:01* | 7.2E-03 | 0.22 | 0.08 | 0.07 | *C\*07:01, B\*13:02, DRB1\*01:01, DRB1\*11:01, C\*07:04, DQB1\*02:02, DRB5\*01:01* |
| *C\*03:04* | 0.02 | 0.12 | 0.05 | 0.23 | *C\*07:01, B\*13:02, DRB1\*01:01, DRB1\*11:01, C\*07:04, DQB1\*02:02, DRB5\*01:01, DRB1\*09:01* |

**Table 2.**
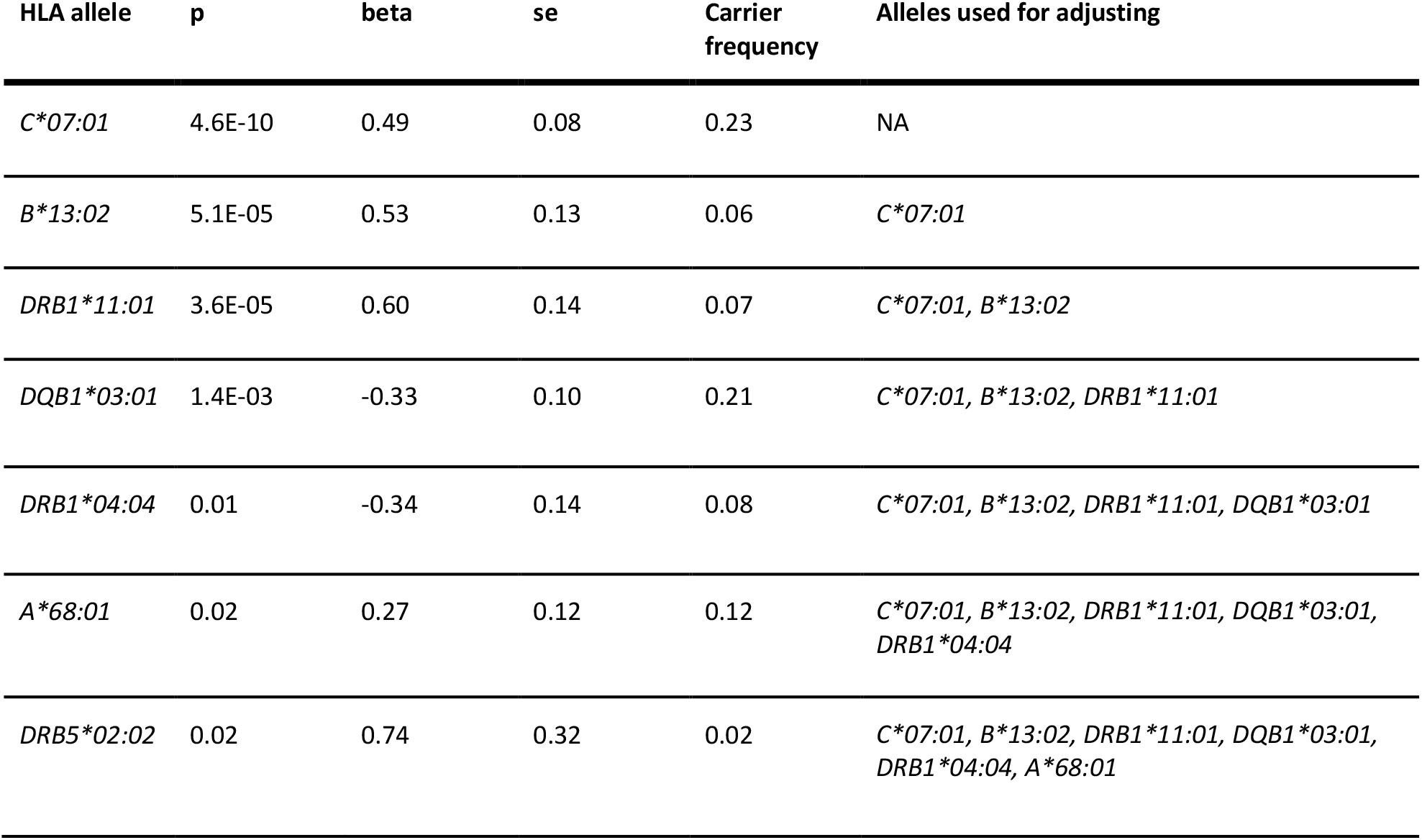
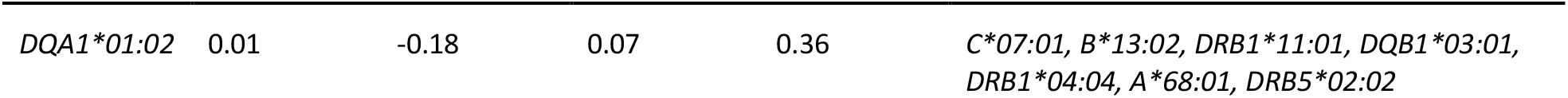
Associations of independent HLA alleles to Puumala virus (PUUV) infection severity were analyzed using fine-mapping and stepwise logistic regression. Individuals with severe PUUV were used as cases and individuals with non-severe PUUV as controls. (se=standard error, carrier frequency = frequency of having at least one copy of allele)

**Figure 3.**
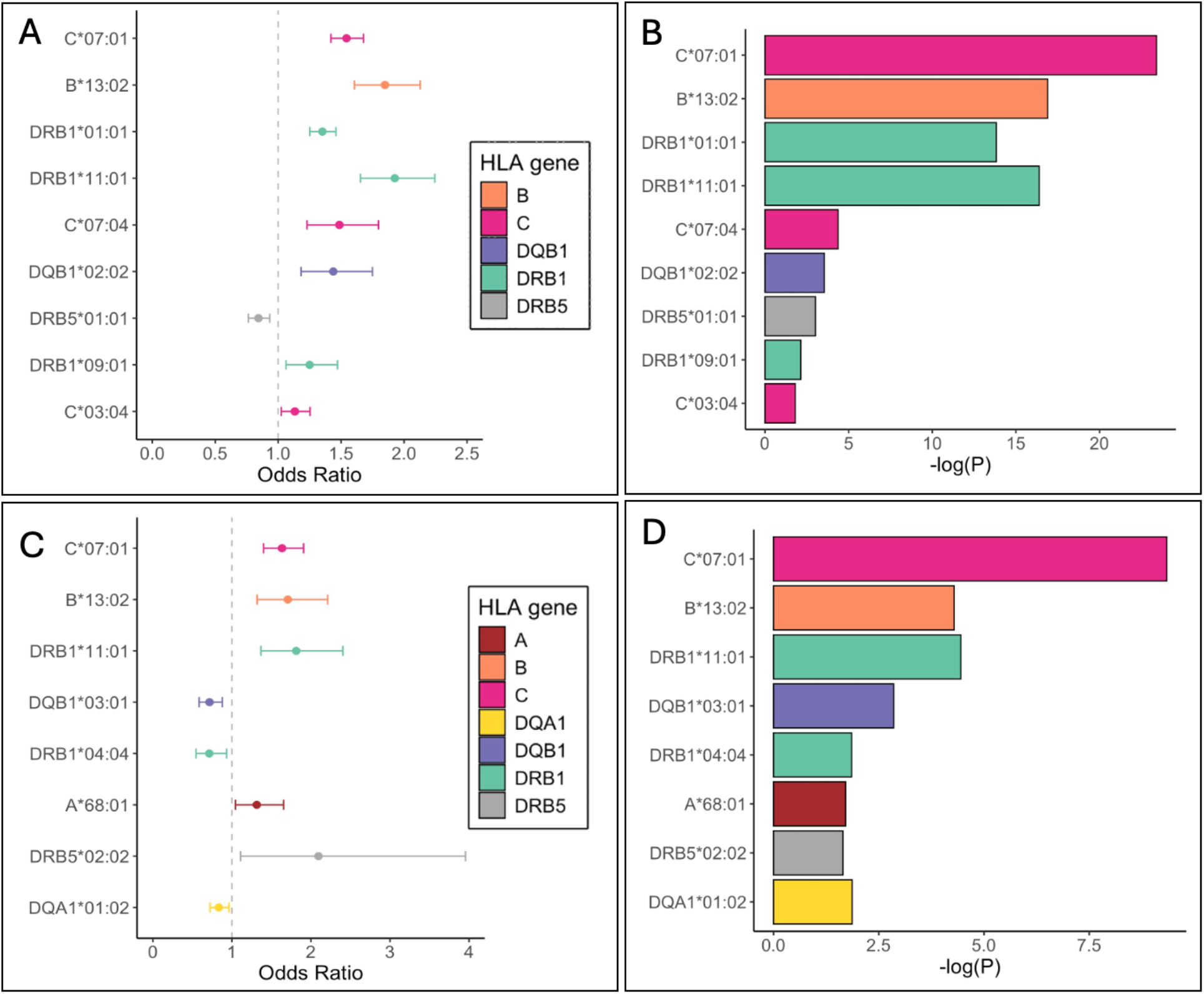
HLA fine-mapping in severe Puumala virus infection. Independent associations of HLA alleles with Puumala virus (PUUV) in FinnGen (P-value < 0.05). Stepwise logistic regression was performed by sequentially adding the most strongly associated HLA allele as a covariate to the multivariate regression analysis until no significant alleles remained. The graphs present alleles in order: the uppermost bar represents the allele that appeared as the most significant before adjusting with HLA alleles. We show odds ratios (A) and p-values (B) from fine-mapping of severe cases using population control and odds ratios (C) and p-values (D) from fine-mapping of severe cases using non-severe cases as controls.

The strongest association for severe PUUV infection were with HLA alleles *C*07:01* (OR = 1.5 [1.5, 1.6], p = 4.0×10^−24^), *B*13:02* (OR = 1.8 [1.7, 2.0], p = 1.2×10^−17^), *DRB1*01:01* (OR = 1.4 [1.3, 1.4], p = 1.5×10^−14^) and *DRB1*11:01* (OR = 1.9 [1.8, 2.1], p = 4.1×10^−17^) in HLA fine-mapping analysis using population control. When assigning severe PUUV infection as cases and non-hospitalized individuals as controls the strongest associations were with *C*07:01* (OR = 1.6 [1.5, 1.8], p = 4.6×10^−10^), *B*13:02* (OR = 1.7 [1.4, 2.0], p = 5.1×10^−05^) and *DRB1*11:01* (OR = 1.8 [1.5, 2.1], p = 3.6×10^−05^).

## Discussion

Here we performed a GWAS to understand host factors and genetic variants that affect the severity of PUUV infection. Utilizing data from the FinnGen cohort with over 2,000 severe cases we identified genetic associations from the HLA locus and *ERAP1* gene to severe PUUV infection. Our lead variant in the *ERAP1* gene was a missense variant changing the amino acid arginine to proline. Furthermore, in the HLA region, we showed independent associations with PUUV severity in both class I and class II regions. With conditional fine-mapping, we further associated the severe disease with several HLA alleles, the most significant of which were *HLA*C:07:01, B*13:02, DRB1*01:01*, and *DRB1*11:01* alleles.

Both the HLA region and *ERAP1* gene are essential for the proper functioning of the adaptive immune response. The HLA region, located in chromosome 6, is exceptionally diverse encoding a vast array of cell surface proteins critical for immune function. These HLA molecules are responsible for presenting antigens to T cells and initiating immune responses against pathogens. Due to their polymorphic nature, variations in HLA genes profoundly impact disease susceptibility and severity in a number of infections. (14)

The *ERAP1* gene, in turn, encodes a protein called endoplasmic reticulum aminopeptidase 1, which acts within the endoplasmic reticulum cleaving diverse viral and bacterial proteins into short peptides, facilitating their recognition by the immune system. Peptides are then transported to the cell surface where they bind to HLA class I molecules, HLA A, HLA B and HLA C proteins, which present viral peptides to the immune system. If the immune system recognizes the peptides as foreign, such as viral or bacterial peptides, it responds by triggering the infected cell to self-destruct. (15)

Previous studies have investigated the potential role of HLA genetic variation in determining susceptibility to PUUV infection and the severity of HFRS. These studies have established a connection between several HLA alleles and haplotypes and the severe course of PUUV. (6-9) Our study is the first GWAS on PUUV infection and we strongly confirm the association to HLA region with two independent variants, one located in HLA class I region in between *HLA-DRB1* and *HLA-DQA1* genes and the other in HLA class II region closest to the *HLA-C* gene. The association at the *ERAP1* locus is a novel finding in the context of PUUV infection yet the gene has previously been linked to various autoimmune disorders such as ankylosing spondylitis and psoriasis, as well as infections including hepatitis C, influenza, and HIV. (16) Based on our findings we hypothesize that variation in the *ERAP1* gene can improve its capability to process PUUV proteins into peptides of suitable length leading to effective presentation on HLA class I molecules, a key step for functional adaptive immune response. In addition, we show that genetic variation in HLA genes shapes the immune response against PUUV.

While the extended HLA region comprises over 200 genes, the majority of association signal is attributed to HLA class I or class II genes and the individual HLA alleles (17). Previous research on PUUV genetics has relied on small case-control studies and mostly focused on the association between specific HLA alleles and haplotypes and the risk of either mild or severe disease. (6-9) However, the implicated alleles have originated from relatively small studies with different alleles associated within each study. In genetic datasets, allelic associations are typically estimated by first imputing classical HLA alleles and then performing regression. Therefore, to understand which genes and alleles contribute to PUUV infection, we performed fine-mapping of HLA alleles, revealing the strongest association at *HLA C*07:01* followed by significant associations with *B*13:02, DRB1*01:01, and DRB1*11:01*. Furthermore, we demonstrated the importance of the *HLA-C*07:01* allele when adjusting our GWAS analysis for the lead allele showing attenuated association in this genomic region. To conclude, our fine-mapping analysis indicates eight independent predisposing and one protective HLA allele for severe disease.

Finally, we also show that the FinnGen sample that covers 10% of the Finnish population gives a representative picture of PUUV infection. We demonstrated that incidence rates vary in cycles of 3-4 years following the changes in vole population density as known from previous research. Peak months for incidences are summer and autumn months. We also showed that men are slightly overrepresented in the cases as well as a higher BMI is associated with the disease. We studied epidemiological associations with previously reported diseases associated with PUUV using multivariate linear regression and Cox models and found statistically significant correlations to thrombocytopenia and acute kidney renal failure, two symptoms commonly linked to PUUV infections.

Our study, the first to utilize large-scale genetic datasets to study PUUV infection, provides a comprehensive view of the disease’s genetic underpinnings. Individual sequencing data and HLA allele information enable us to employ genetic tools and methods for reliable, unbiased analysis. PUUV infections are prevalent in Finland, with diagnoses verified through laboratory testing, which provides ideal conditions for investigating PUUV genetics. Nonetheless, our study has some limitations. Our analyses were exclusively performed within the FinnGen study and thus only contained individuals of Finnish ancestry. For this reason, we cannot say whether the results are generalisable to other ethnicities than Europeans. Additionally, in Finland, Puumala viruses cause only HFRS, whereas in other countries Hantavirus Pulmonary syndrome is a common form of the infection as well. Thus, our results are specific to HFRS, and corresponding studies for HPS would be highly beneficial. Lastly, our epidemiological analysis lacks sufficient sample size, as FinnGen covers only about 10% of Finns, and we were unable to replicate some previous findings identified using nationwide registries. It is also noteworthy that the FinnGen dataset is enriched for disease endpoints due to the relatively high median age of participants and the significant proportion of hospital-based recruitment (18).

Our genetic study on PUUV implicates peptide processing and presentation relevant to severe PUUV infection. *ERAP1* gene polymorphism and alleles from both HLA class I and II are associated with the severe disease, shedding light on disease mechanisms and immune defense against the Puumala virus. Overall, these findings highlight the well-known association between HLA molecules and infections, suggesting that both HLA class I and class II variation contribute to PUUV infection and finally implicate the role of ERAP1, a known immune mediator, also in PUUV infection.

## Methods

### Study cohort

***FinnGen*** is a population-based public-private population cohort established in 2017. (18) The study combines genetic data with electronic health record data, including International Classification of Diseases (ICD) codes spanning an individual’s entire lifespan, derived from primary care registers, hospital inpatient and outpatient visits, drug prescriptions and several other registries. The project aims to improve understanding of the genetic etiology of diseases and disorders potentially leading to drug development. For this study, we leveraged data from FinnGen’s data freeze 12, which includes 520,210 participants.

To identify Puumala virus cases from the cohort ICD10 code A98.5 for Hemorrhagic Fever with Renal Syndrome has been used. Data was derived from hospital and primary care registers. The severe cases were classified as those requiring hospitalization whereas the non-severe cases were not hospitalized.

Total number of all diagnosed PUUV cases in Finland were obtained from the National Registry for Infectious DIseases at the National Institute for Health and Welfare, from publicly available data (18). All laboratories notify new diagnoses of communicable diseases there.

### FinnGen ethics statement

Patients and control subjects in FinnGen provided informed consent for biobank research, based on the Finnish Biobank Act. Alternatively, separate research cohorts, collected prior the Finnish Biobank Act came into effect (in September 2013) and start of FinnGen (August 2017), were collected based on study-specific consents and later transferred to the Finnish biobanks after approval by Fimea (Finnish Medicines Agency), the National Supervisory Authority for Welfare and Health. Recruitment protocols followed the biobank protocols approved by Fimea. The Coordinating Ethics Committee of the Hospital District of Helsinki and Uusimaa (HUS) statement number for the FinnGen study is Nr HUS/990/2017.

The FinnGen study is approved by Finnish Institute for Health and Welfare (permit numbers: THL/2031/6.02.00/2017, THL/1101/5.05.00/2017, THL/341/6.02.00/2018, THL/2222/6.02.00/2018, THL/283/6.02.00/2019, THL/1721/5.05.00/2019 and THL/1524/5.05.00/2020), Digital and population data service agency (permit numbers: VRK43431/2017-3, VRK/6909/2018-3, VRK/4415/2019-3), the Social Insurance Institution (permit numbers: KELA 58/522/2017, KELA 131/522/2018, KELA 70/522/2019, KELA 98/522/2019, KELA 134/522/2019, KELA 138/522/2019, KELA 2/522/2020, KELA 16/522/2020), Findata permit numbers THL/2364/14.02/2020, THL/4055/14.06.00/2020, THL/3433/14.06.00/2020, THL/4432/14.06/2020, THL/5189/14.06/2020,THL/5894/14.06.00/2020, THL/6619/14.06.00/2020, THL/209/14.06.00/2021, THL/688/14.06.00/2021, THL/1284/14.06.00/2021, THL/1965/14.06.00/2021, THL/5546/14.02.00/2020, THL/2658/14.06.00/2021, THL/4235/14.06.00/2021, Statistics Finland (permit numbers: TK-53-1041-17 and TK/143/07.03.00/2020 (earlier TK-53-90-20) TK/1735/07.03.00/2021, TK/3112/07.03.00/2021) and Finnish Registry for Kidney Diseases permission/extract from the meeting minutes on 4th July 2019.

The Biobank Access Decisions for FinnGen samples and data utilized in FinnGen Data Freeze 10 include: THL Biobank BB2017_55, BB2017_111, BB2018_19, BB_2018_34, BB_2018_67, BB2018_71, BB2019_7, BB2019_8, BB2019_26, BB2020_1, BB2021_65, Finnish Red Cross Blood Service Biobank 7.12.2017, Helsinki Biobank HUS/359/2017, HUS/248/2020, HUS/150/2022 § 12, §13, §14, §15, §16, §17, §18, and §23, Auria Biobank AB17-5154 and amendment #1 (August 17 2020) and amendments BB_2021-0140, BB_2021-0156 (August 26 2021, Feb 2 2022), BB_2021-0169, BB_2021-0179, BB_2021-0161, AB20-5926 and amendment #1 (April 23 2020)and it’s modification (Sep 22 2021), Biobank Borealis of Northern Finland_2017_1013, 2021_5010, 2021_5018, 2021_5015, 2021_5023, 2021_5017, 2022_6001, Biobank of Eastern Finland 1186/2018 and amendment 22 § /2020, 53§/2021, 13§/2022, 14§/2022, 15§/2022, Finnish Clinical Biobank Tampere MH0004 and amendments (21.02.2020 & 06.10.2020), §8/2021, §9/2022, §10/2022, §12/2022, §20/2022, §21/2022, §22/2022, §23/2022, Central Finland Biobank 1-2017, and Terveystalo Biobank STB 2018001 and amendment 25th Aug 2020, Finnish Hematological Registry and Clinical Biobank decision 18th June 2021, Arctic biobank P0844: ARC_2021_1001.

### Genotyping and quality control

The samples of FinnGen were genotyped using Illumina (Illumina) and Affymetrix arrays (Thermo Fisher Scientific). The array consists of 735,145 probes that capture 655,973 variants encompassing core backbone variants essential for imputation, rare coding variants that are enriched in the Finnish population, as well as variants associated with KIR and HLA haplotypes. Additionally, the array includes markers specific to certain diseases and pharmacogenomic markers. (19)

To ensure data integrity, genotyping information from prior chip platforms and reference genome builds was lifted over to build 38 (GRCh38/hg38). Rigorous sample-wise quality control measures were implemented, including the exclusion of individuals with discrepancies between genetically inferred sex and reported sex in registries, high genotype missingness (>5%), and excess heterozygosity (±4 standard deviations). Variant-level quality control involved filtering out variants with high missingness (>2%), low Hardy–Weinberg equilibrium (P < 1 × 10^−6^), and a minor allele count < 3. (19)

For further refinement, chip-genotyped samples underwent pre-phasing with Eagle 2.3.5, followed by imputation using the Finnish-specific SISu v4 imputation reference panel. Post-imputation quality control criteria included the exclusion of variants with an INFO score < 0.7. (19)

### HLA imputation

HLA imputation in FinnGen was performed for *HLA A, HLA B, HLA C, HLA DRB1, HLA DQA1, HLA DQB1, HLA DPB1, HLA DRB3, HLA DRB4* and *HLA DRB5* using R library HIBAG (HLA genotype imputation with attribute bagging), as described by Ritari et al. (20). The HLA imputation of FinnGen data is based on a set of SNPs directly genotyped on the FinnGen array. A Finnish reference panel, genotyped at clinical grade accuracy (4-digits, amino acid), was used.

### Genetic analyses

GWAS in FinnGen was conducted using the REGENIE pipeline (https://github.com/FINNGEN/regenie-pipelines) adjusting for age, sex, chip, batch and ten first principal components. The ICD-10 code for hemorrhagic fever with renal syndrome (A98.5), was used to identify individuals with PUUV infection. The cases were further divided to severe and non-severe based on their need for hospitalization. GWAS analyses were performed for severe PUUV infection versus the remainder of the FinnGen participants and for severe PUUV infection versus non-severe PUUV infection. Additional GWAS for sensitivity assessments were performed between PUUV against population control and non-severe PUUV against population control (Table S5). Manhattan plots were created using R version 4.0.1 (packages: qqman (21) and RColorBrewer). Miami plots were created using the Miamiplot R package (22).

Association testing for HLA variants was conducted using multivariate logistic regression to elucidate the relationships between individual HLA alleles and PUUV infection. Multivariate logistic regression analysis was adjusted for age at death or the end of follow-up, sex, and the first 10 genetic principal components accounting for population structure. Multivariate logistic regression was performed in a stepwise manner by sequentially adding the most strongly associated HLA allele as a covariate to the analysis. This iterative process was repeated until no significant (p<0.05) alleles remained. The multivariate logistic regression analyses were conducted using R 4.0.1 and utilized the packages data.table (23), dplyr (24), and tidyverse (25).

### Epidemiological analyses

We conducted logistic regression analysis to explore associations between PUUV and thrombocytopenia (ICD-10: D69.6, D69.59), acute renal failure (ICD-10: N17, N18, N19, N08.1, N16.0, N29.1), thrombosis (ICD-10: I80.1, I80.2, I80.3, I80.8, I80.9, I81.9, I82.2, I82.3, I82.8, I82.9), lymphoid malignancies (ICD-10: C81-C96) and hypopituitarism (ICD-10: E23.0) in FinnGen (Table S1). The model was adjusted for age at death or end of follow-up, sex and the first 10 genetic principal components. Additionally, we constructed a model that was further adjusted for BMI in addition to the covariates mentioned above.

To evaluate the temporal aspect, we employed the Cox proportional hazards model with age as the timescale and sex as a covariate. Prevalent cases, meaning the individuals who had obtained the studied conditions prior to PUUV infection, were excluded from the analysis. (26)

## Data Availability

All data produced in the present study are available upon reasonable request to the authors.

## Supplementary material

Supplemental material includes one Microsoft Excel file (with 3 tables) and one Microsoft Word file (with Supplementary Figures S1-S5 and Supplementary Tables S1-S5).

## Author contributions

HH, SS and HMO designed the study and both analyzed and verified the underlying data. HMO, MK and AMFC provided mentorship and intellectual contributions. HH, SS and HMO wrote the manuscript. SS, AT, SJ, HMO, MK and AMFC revised the manuscript. All contributing authors have read and approved the final version of this manuscript.

## Data and Code Availability

Summary level data will be shared in the GWAS catalog at the time of publication. Other (non-individual level) data and code used in this study are available upon reasonable request. To access the individual level data in FinnGen, please visit the Fingenious portal (https://site.fingenious.fi/en/) hosted by the Finnish Biobank Cooperative.

## Acknowledgments

We want to acknowledge the participants and investigators of the FinnGen study. The FinnGen project is funded by two grants from Business Finland (HUS 4685/31/2016 and UH 4386/31/2016) and the following industry partners: AbbVie Inc., AstraZeneca UK Ltd, Biogen MA Inc., Bristol Myers Squibb (and Celgene Corporation & Celgene International II Sàrl), Genentech Inc., Merck Sharp & Dohme LCC, Pfizer Inc., GlaxoSmithKline Intellectual Property Development Ltd., Sanofi US Services Inc., Maze Therapeutics Inc., Janssen Biotech Inc, Novartis Pharma AG, and Boehringer Ingelheim International GmbH. Following biobanks are acknowledged for delivering biobank samples to FinnGen: Auria Biobank (www.auria.fi/biopankki), THL Biobank (www.thl.fi/biobank), Helsinki Biobank (www.helsinginbiopankki.fi), Biobank Borealis of Northern Finland (https://www.ppshp.fi/Tutkimus-ja-opetus/Biopankki/Pages/Biobank-Borealis-briefly-in-English.aspx), Finnish Clinical Biobank Tampere (www.tays.fi/en-US/Research_and_development/Finnish_Clinical_Biobank_Tampere), Biobank of Eastern Finland (www.ita-suomenbiopankki.fi/en), Central Finland Biobank (www.ksshp.fi/fi-FI/Potilaalle/Biopankki), Finnish Red Cross Blood Service Biobank (www.veripalvelu.fi/verenluovutus/biopankkitoiminta), Terveystalo Biobank (www.terveystalo.com/fi/Yritystietoa/Terveystalo-Biopankki/Biopankki/) and Arctic Biobank (https://www.oulu.fi/en/university/faculties-and-units/faculty-medicine/northern-finland-birth-cohorts-and-arctic-biobank). All Finnish Biobanks are members of BBMRI.fi infrastructure (www.bbmri.fi). Finnish Biobank Cooperative -FINBB (https://finbb.fi/) is the coordinator of BBMRI-ERIC operations in Finland. The Finnish biobank data can be accessed through the Fingenious® services (https://site.fingenious.fi/en/) managed by FINBB.

H.H. received funding for this project from Finland’s Doctoral Education Pilot project and A.T. received support for this work from the Instrumentarium Science Foundation (230041).

