## Supplementary material for "Genetic analysis implicates *ERAP1* and HLA as risk factors for severe Puumala virus infection"

### Supplementary word

#### 1. Epidemiological analyses

Previous research studies and case reports have documented an elevated occurrence of thrombocytopenia, acute renal failure, thrombosis, lymphoid malignancies, and hypopituitarism subsequent to PUUV infection. We employed multivariate logistic regression and Cox proportional hazard models to study the associations of the previously reported conditions with PUUV. ICD codes used to extract the cases and number of cases in the PUUV case and control subset are reported in Table S1. PUUV cases were identified with ICD10 code A98.5 Hemorrhagic fever with renal syndrome.

**Table S1.** ICD codes used to define conditions previously linked to PUUV infections and case numbers in PUUV infected and controls in FinnGen.

| | ICD10 code | Cases in PUUV cases | Cases in PUUV controls |
|---|---|---|---|
| **Thrombocytopenia** | D69.6, D69.59 | 53 | 3519 |
| **Acute renal failure** | N17, N18, N19, N08.1, N16.0, N29.1 | 497 | 3088 |
| **Thrombosis** | I80.1, I80.2, I80.3, I80.8, I80.9, I81.9, I82.2, I82.3, I82.8, I82.9 | 181 | 3519 |
| **Lymphoid malignancies** | C81-C96 | 91 | 3483 |
| **Hypopituitarism** | E23.0 | 39 | 3533 |

We conducted logistic regression analysis to explore associations between PUUV and thrombocytopenia, acute renal failure, thrombosis, lymphoid malignancies and hypopituitarism in FinnGen. The model was adjusted for baseline age, sex and the first 10 genetic principal components. Additionally, we constructed a model that was further adjusted for BMI, in addition to the covariates mentioned above. (Table S2)

**Table S2.** Logistic regression model results for the association between PUUV and thrombocytopenia, acute renal failure, thrombosis, lymphoid malignancies and hypopituitarism in FinnGen.

| | **OR [95 % CI] (not BMI adjusted)** | **p-value (not BMI adjusted)** | **OR [95 % CI] (BMI adjusted)** | **p-value (BMI adjusted)** |
|---|---|---|---|---|
| **Thrombocytopenia** | 2.58 [1.99, 3.35] | $1.23x10^{-11}$ | 2.46 [1.84, 3.30] | $5.14x10^{-8}$ |
| **Acute renal failure** | 1.32 [1.28, 1.35] | $4.00x10^{-8}$ | 1.31 [1.27, 1.35] | $4.33x10^{-6}$ |
| **Hypopituitarism** | 1.77 [1.48, 2.12] | $4.47x10^{-4}$ | 1.69 [1.39,2.05] | $6.50x10^{-3}$ |
| **Lymphoid malignancies** | 1.21 [1.16, 1.26] | 0.077 | 1.33 [1.24, 1.42] | 0.019 |
| **Thrombosis** | 1.22 [1.19, 1.26] | $9.22x10^{-3}$ | 1.24 [1.19, 1.28] | 0.018 |

To evaluate the temporal aspect, we employed the Cox proportional hazard model with age as the timescale and sex as a covariate. Prevalent cases were excluded from the model. (Table S3)

**Table S3.** COX proportional hazard model results for the association between PUUV and thrombocytopenia, acute renal failure, thrombosis, lymphoid malignancies and hypopituitarism in FinnGen.

| | **HR [95 % CI]** | **p-value** |
|---|---|---|
| **Acute renal failure** | 1.22 [1.10, 1.34] | $9.87x10^{-5}$ |
| **Thrombocytopenia** | 1.86 [1.34, 2.58] | $2.22x10^{-4}$ |
| **Hypopituitarism** | 1.12 [0.76, 1.65] | 0.56 |
| **Lymphoid malignancies** | 0.85 [0.65, 1.10] | 0.21 |
| **Thrombosis** | 0.76 [0.63, 0.92] | $4.58x10^{-3}$ |

#### 2. GWAS analyses

GWAS in FinnGen was conducted using the REGENIE pipeline adjusting with age, sex, chip, batch and ten first principal components. Using the ICD10 code for hemorrhagic fever with renal syndrome (A98.5), we identified 3,650 PUUV cases. They were classified as severe and non-severe disease according to the need for hospitalization treatment. 2,227 of the patients were admitted to the hospital and classified as severe whereas the remaining 1,423 cases were non-severe.

We ran GWAS analyses for two phenotypes:

1. Severe PUUV vs population control (Figure S1 and Figure S3A-C)
2. Severe PUUV vs non-severe PUUV (Figure 2 and Figure 3D)

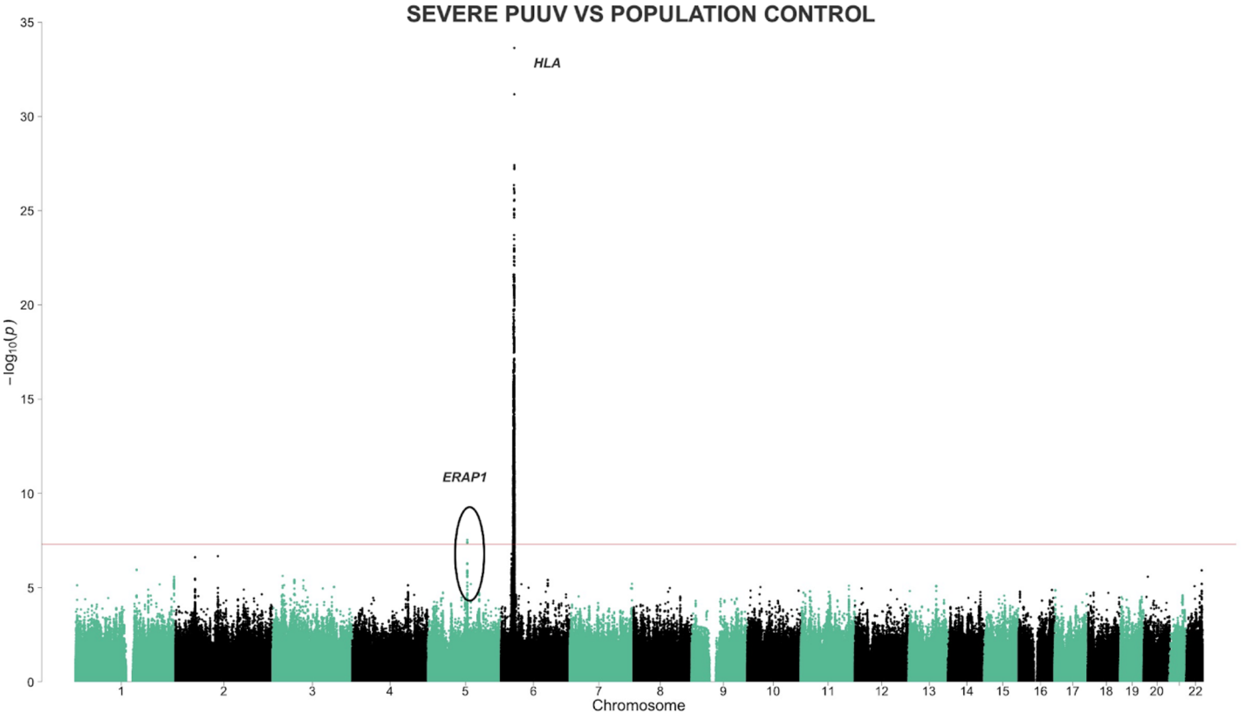

**Figure S1. Manhattan plot for Severe PUUV vs population control (=remaining individuals) in FinnGen.**

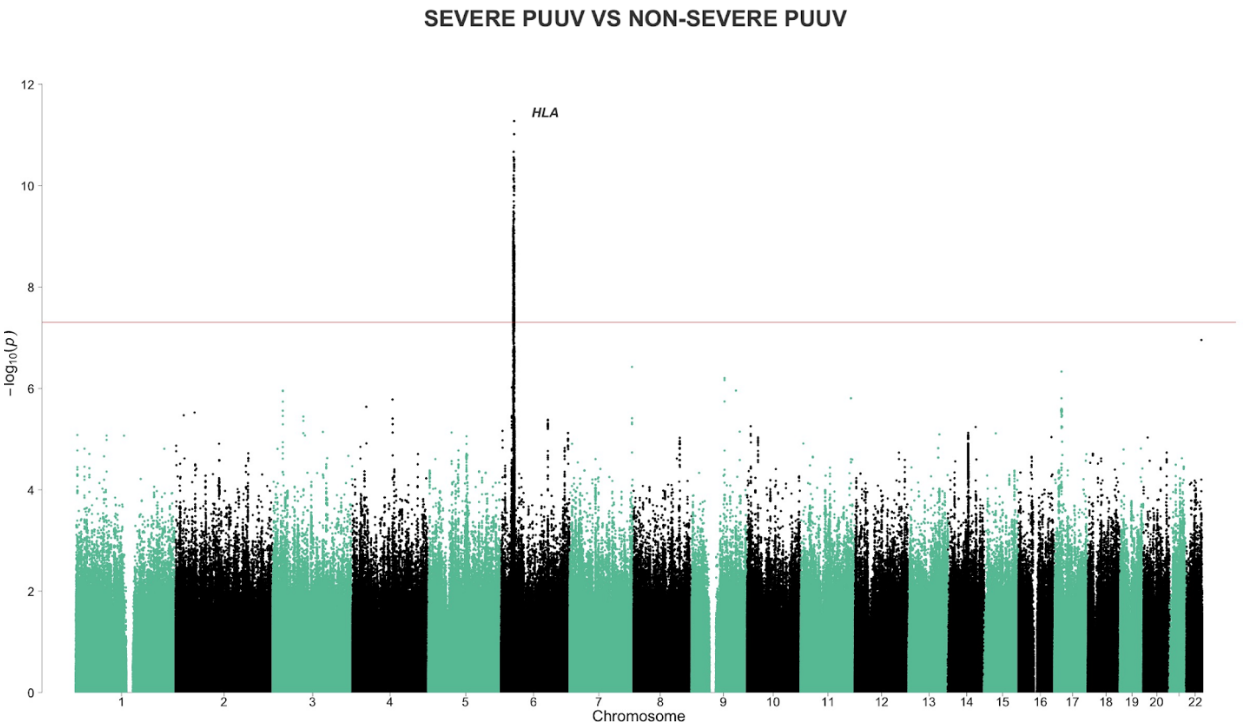

**Figure S2. Manhattan plot for Severe PUUV vs Non-Severe PUUV in FinnGen.**

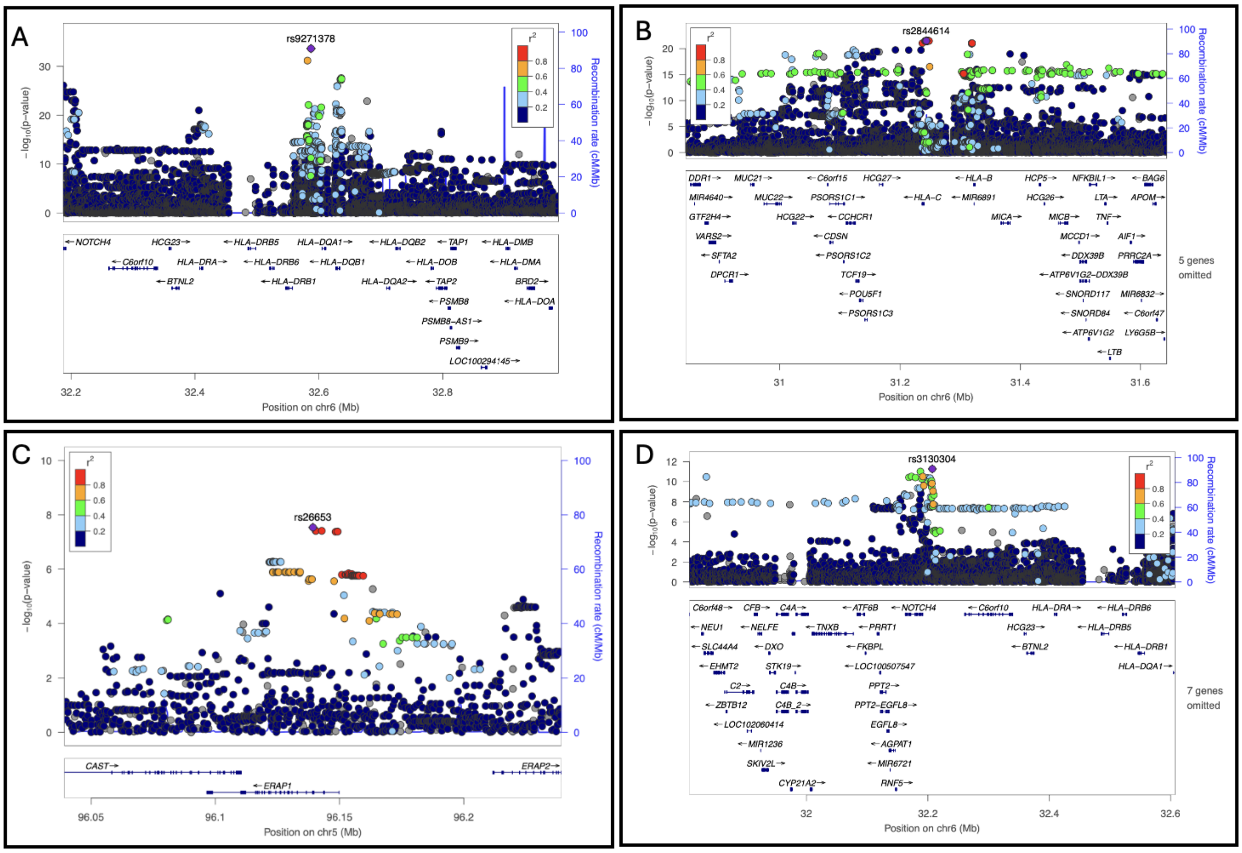

**Figure S3. Locus zoom plots for severe PUUV infection lead variants.** A. Locus zoom plot for Severe PUUV vs population control HLA lead variant rs9271378. B. Locus zoom plot for Severe PUUV vs population control second independent HLA variant rs2844614. C. Locus zoom plot for Severe PUUV vs population control *ERAP1* gene lead variant rs26653. D. Locus zoom plot for Severe PUUV vs Non-Severe PUUV HLA lead variant rs3130304.

The genome-wide significant lead signals are listed in Table S4.

**Table S4.** Genome-wide significant lead variants from GWAS analyses for Severe PUUV vs population control and Severe PUUV vs Non-Severe PUUV in FinnGen.

| Dataset | chr | position | ref | alt | rsid | pval | beta | af_alt |
|---|---|---|---|---|---|---|---|---|
| **Severe PUUV vs Population Control** | 6 | 32619523 | A | G | rs9271378 | $2.30 \times 10^{-34}$ | 0.36 | 0.41 |
| | 5 | 96803547 | C | G | rs26653 | $2.93 \times 10^{-8}$ | -0.18 | 0.71 |
| **Severe PUUV vs Non-Severe PUUV** | 6 | 32239404 | G | A | rs3130304 | $5.34 \times 10^{-12}$ | 0.46 | 0.19 |

Additionally, the signal in the *ERAP1* gene in chromosome 5 showed up in Severe PUUV vs Non-Severe PUUV GWAS analysis, although below the genome-wide significance threshold (rs26653, $p = 8.3 \times 10^{-5}$, beta = -0.22).

To evaluate potential additional independent GWAS signals in the HLA region, we adjusted both GWAS analyses with their lead variant. Conditional analysis of Severe PUUV vs Population control revealed an additional independent genome-wide significant signal with lead variant rs2844614 ($P = 9.97 \times 10^{-14}$, beta = 0.31). This variant was located at the HLA class I region closest to the HLA C allele (see Figure 3B). In Severe PUUV vs non-severe PUUV the conditioning with the lead variant explained the signal quite well leaving no genome-wide significant associations. The p-value of the lead variant from Severe PUUV vs Population control GWAS analysis, rs2844614, was $p = 2.54 \times 10^{-4}$, which is significant at the HLA level.

### 1. Sensitivity analysis

Additional sensitivity analyses were performed to study whether the associations are linked more to disease severity than susceptibility. We conducted GWAS analyses for:

1. PUUV against population control
2. Non-severe PUUV against population control

We identified HLA association with the same lead variant from PUUV case vs control GWAS. The smaller P-value and effect size indicate that association is more related to disease severity than susceptibility. The Non-severe PUUV vs population control GWAS identified no significant associations and the p-value of rs9271378 was $1.95 \times 10^{-3}$ supporting the finding of the HLA association being related to severe disease. (Table S5)

**Table S5.** GWAS association signals with PUUV and Non-Severe PUUV in FinnGen. (*no significant associations)

| Dataset | HLA lead variant | HLA lead pval | HLA lead BETA | af_alt |
|---|---|---|---|---|
| **PUUV vs No PUUV** | rs9271378 | $3.03 \times 10^{-30}$ | 0.27 | 0.41 |
| **Non-severe PUUV vs Population Control*** | rs9271378 | $1.95 \times 10^{-3}$ | 0.12 | 0.41 |

#### 2. Fine mapping

Association testing for HLA variants was conducted using multivariate logistic regression to elucidate the relationships between individual HLA alleles and PUUV infection. Multivariate logistic regression analysis was adjusted for age at death or at the end of follow-up, sex, and the first 10 genetic principal components accounting for population structure. The analysis was performed in stepwise manner by sequentially adding the most strongly associated HLA allele as a covariate to the analysis. This iterative process was repeated until no significant alleles remained in the analysis.

Only HLA alleles with allele frequency > 1 % were taken into account in the analysis.

The HLA allele frequencies in FinnGen are presented in Supplementary Excel together with all signals from the first round of fine mapping and the independent lead variants. The analyses were performed for Severe PUUV vs population control and Severe PUUV vs Non-severe PUUV cohorts.

The strongest association was with the *HLA-C*07:01*. The second independent lead variant from our GWAS was located closest to HLA C supporting our findings of the relevant HLA alleles. We further assessed the degree of explanation of this allele by adjusting GWAS analysis with it. Severe PUUV vs population control GWAS conditioned with the *HLA-C*07:01* had a smaller lead variant p-value (rs9282211, $P = 1.0 \times 10^{-28}$) compared to the non-adjusted model as well as Severe PUUV vs Non-severe PUUV GWAS (rs70993814, $P = 2.6 \times 10^{-7}$). This indicates that the *HLA-C*07:01* allele is explaining some but not all variation in the disease status. The result aligns with our findings of several independently associated HLA alleles.
